# Associations between initial treatments for acute low back pain and opioid use disorder and overdose risk in Medicaid patients

**DOI:** 10.64898/2026.06.05.26355003

**Authors:** Lisa V. Doan, Anton M. Hung, Mark Olfson, Nicholas T. Williams, Kara E. Rudolph

**Author notes:** **Corresponding author contact information:** Kara E. Rudolph, 722 West 168th St, 16th floor, New York, NY 10032.

## Abstract

**Introduction:** Acute low back pain is a leading cause of disability worldwide. Clinical guidelines recommend non-pharmacological therapies as first-line treatment and advise caution with opioid prescribing. However pharmacological therapies, including opioids and gabapentinoids, remain commonly used. The comparative risks of subsequent opioid use disorder (OUD) and overdose diagnosis associated with initial treatment modality in large, real-world populations is not well characterized. We estimated the incidence of new-onset OUD and overdose diagnosis among opioid-naïve, Medicaid-insured adults with newly diagnosed acute low back pain and estimated the association between initial treatment modalities and subsequent OUD and overdose diagnosis risk.

**Methods:** We conducted a retrospective cohort study using Medicaid T-MSIS Analytic files from 25 states (2016-2019). We identified opioid-naïve adults with a new diagnosis of acute low back pain who initiated pharmacologic or non-pharmacologic treatment within 1 month of diagnosis. The primary outcome was incident OUD and overdose diagnosis (based on diagnosis codes in claims) during follow-up. Associations between initial treatment modality and OUD and overdose diagnosis risk were estimated using a non-parametric, doubly robust estimator to adjust for measured confounding.

**Results:** The cohort included 525,002 opioid-naïve adults initiating treatment for low back pain. The cumulative incidence of OUD and overdose diagnosis was 1.5% and 2.4% at 7 and 13 months, respectively. Compared to non-use, use of gabapentinoids during the first month of treatment was associated with the highest relative risk (increasing risk by 130.1%, 95% confidence interval (CI): 117.8%, 142.3%), the second-highest relative risk was estimated for higher-dose opioids, defined as > 50 daily Morphine Milligram Equivalents (MME) (118.1%, 95% CI: 99.2%, 137.0%). Lower-dose, short-duration opioids (≤ 50 MME, ≤ 7 days) were also associated with elevated risk, though substantially smaller in magnitude (20.8%, 95% CI: 13.8%, 27.9%). In contrast, non-pharmacologic, non-interventional therapies were associated with reduced OUD and overdose diagnosis risk, with physical therapy demonstrating the largest relative reduction of 34.0% (95% CI: −40.9%, −27.1%).

**Discussion:** In opioid-naive Medicaid patients with acute low back pain, initial non-pharmacologic treatment was associated with reduced OUD and overdose diagnosis risk. Gabapentinoids and opioids were each associated with increased risk; for opioids, the degree of risk increased with higher doses and durations. These results support guideline recommendations favoring non-pharmacologic treatment as first-line therapy and indicate the importance of cautious prescribing when pharmacologic treatment is considered.

## Introduction

Acute low back pain (aLBP) is one of the most common reasons adults seek medical care.^[1]^ The one-year prevalence of aLBP is estimated to be as high as 38% with a lifetime prevalence of approximately 40%.^[2]^ Clinical practice guidelines from the American College of Physicians for the management of aLBP recommend non-pharmacologic therapies as first-line treatment, and suggest nonsteroidal anti-inflammatory drugs (NSAIDs) or skeletal muscle relaxants if pharmacological therapy is considered.^[3]^ Similarly, the 2022 Centers for Disease Control and Prevention (CDC) Clinical Practice Guideline for Prescribing Opioids for Pain recommends nonopioid pharmacologic and non-pharmacologic therapies as first-line treatment for acute pain, including aLBP.^[4]^

Despite these recommendations, guideline-discordant care remains common. In a retrospective analysis of 2017 US outpatient administrative claims data, 13.7% of privately insured visits and 19% of Medicaid visits for aLBP had an associated opioid prescription.^[5]^ In a separate Medicaid claims analysis from 2018, 12.1% of patients received an opioid prescription as the initial aLBP treatment, 57.8% received a nonopioid prescription, and 25.1% received noninvasive, non-pharmacologic treatment as first aLBP treatment modalities. ^[6]^ Guideline-discordant care for aLBP has been associated with adverse outcomes, including progression to chronic LBP, increased disability, and higher health care costs.^[7,8]^ In addition, use of higher-risk medications may increase the likelihood of opioid-related harms. Among Medicare patients with aLBP, those who were prescribed opioids and/or gabapentinoids had an almost twofold increased odds of overdose-related hospitalization within 90 days of aLBP diagnosis.^[9]^ In a prior study of Medicaid patients with chronic pain, opioids prescribed with gabapentinoids or benzodiazepines were significantly associated with increased risk of opioid use disorder (OUD), while physical therapy was associated with decreased risk.^[10]^

Given the high prevalence of aLBP, it is important to understand how initial treatment decisions influence subsequent risk of new-onset diagnoses of OUD and overdose for at least two reasons. First, quantifying risks associated with opioids and other potentially guideline-discordant therapies could reinforce initiatives to promote guideline-concordant care. Second, important differences in risk may exist across the range of pharmacologic and non-pharmacologic therapies for aLBP, including non-opioid treatments that may be perceived as safe options. For example, emerging evidence suggests substantial risks of opioid-related adverse outcomes associated with gabapentoid prescriptions even in the absence of an overlapping opioid prescription.^[11,12]^

In this study, we examined an opioid-naïve adult Medicaid population with newly diagnosed aLBP and estimated the association between initial treatment modality and subsequent risk of new-onset OUD and opioid overdose diagnoses. We hypothesized that non-pharmacologic treatments would be associated with lower risk of OUD and overdose diagnoses while use of medications such as opioids and gabapentinoids would be associated with higher risk.

## Methods

### Study Sample

Data for this study came from the Transformed Medicaid Statistical Information System (T-MSIS) Analytic Files (TAF); including the Demographic and Eligibility, Other Services, Inpatient, and Pharmacy files from 2016-2019. We included data from the 25 states that adopted Affordable Care Act Medicaid expansion by 2014 (AR, AZ, CA, CO, CT, DE, HI, IA, IL, KY, MA, MI, MN, ND, NH, NJ, NM, NV, NY, OH, OR, RI, VT, WA, WV),^[13]^ excluding MD due to unreliable diagnosis code data.^[14]^ Additional details of the data extraction are given in Section S1 of the Supplement.

We created a cohort of Medicaid beneficiaries aged 19-63 years with newly diagnosed aLBP, defined as an International Classification of Diseases (ICD)-10 diagnosis of low back pain with no prior low-back pain diagnoses during a 6-month washout period. ICD-10 diagnosis codes for low-back pain are provided in Table S1. Patients were required to initiate a pharmacologic or non-pharmacologic pain treatment within 1 month of diagnosis. The date of first treatment initiation was designated as day 0 and marked the start of follow-up. We also excluded individuals who were diagnosed with any other pain condition during the 3 months preceding day 0, and excluded those who had their initial pain diagnosis or therapy in an inpatient or residential setting. We excluded patients for not being continuously enrolled in Medicaid during this washout period and for having any history of: OUD diagnosis, overdose, medication for treating OUD, opioid prescription, pregnancy, dual eligibility for Medicaid and Medicare, cancer, or institutionalization. All cohort eligibility criteria were evaluated during a 6-month washout period prior to day 0 (Figure 1).

**Figure 1:**
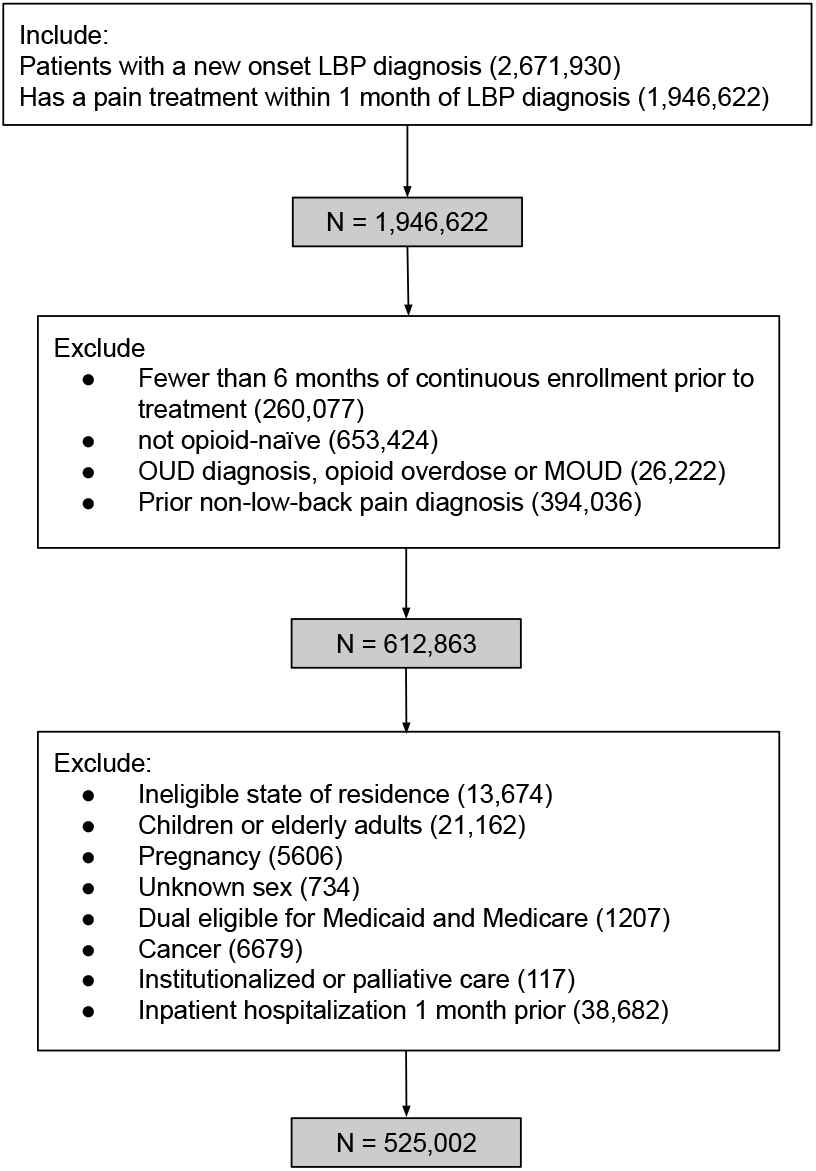
Flowchart of cohort inclusion and exclusion criteria

**Figure 2:**
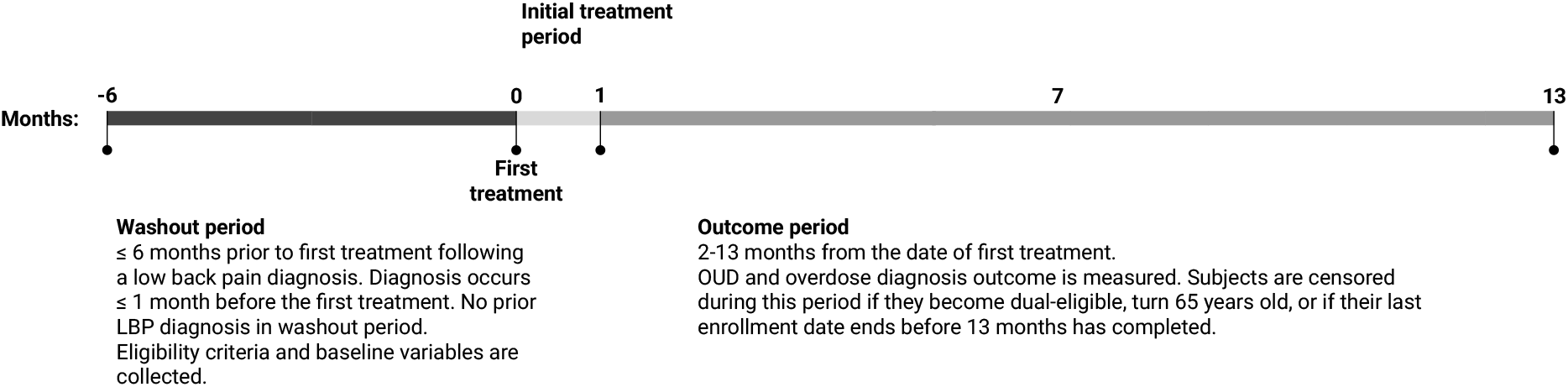
Study measure characterization timeline

Patients were followed for 13 months from day 0. Initial treatment exposure was defined during the first 30 days from day 0. Outcomes were assessed after day 30 and through month 13 to ensure temporal separation between exposure and outcome. Individuals were censored at loss of Medicaid enrollment, Medicare eligibility, death (if recorded), or December 31, 2019.

### Covariates

We characterized each patient’s baseline covariates during the 6-month washout period. The demographic covariates included age, sex, race and ethnicity, primary language, marital status, household size, veteran status, income category, whether or not they received Temporary Assistance for Needy Families (TANF) benefits or Disability Insurance (SSDI) benefits. Clinical covariates included alcohol use disorder, other substance use disorder, and any inpatient or outpatient diagnosis of psychiatric disorders including diagnoses of bipolar disorder, anxiety disorder, major depressive disorder, attention-deficit/hyperactivity disorder, or other psychiatric disorder, and whether they received any mental health counseling. Health system utilization measures included the number of inpatient hospitalizations, outpatient visits, and emergency department visits during the washout period.

### Exposures

The exposure was the set of initial treatments received for aLBP. The initial treatment period began on day 0 and ended at day 30. In sensitivity analyses, we defined the initial treatment period as ending: 1) at the first 30-day gap in pain treatment or at the end of 3 months, whichever occurred first; and in a second sensitivity analysis 2) at the first 7-day gap in pain treatment or at the end of 3 months, whichever occurred first.

Pharmacologic treatments included acetaminophen, anti-inflammatories, benzodiazepines, gabapentinoids, duloxetine, muscle relaxants, steroids, and opioid analgesics (excluding opioid formulations indicated for OUD treatment).^[15–17]^ Opioid prescriptions were categorized based on days supply and maximum daily milligrams of morphine equivalents (MME): ≤ 7 days supply and ≤ 50 MME; > 7 days supply and ≤ 50 MME; and > 50 MME. Opioid doses > 50 MME are associated with higher risk of overdose compared to lower dosages, with no evidence of clinicial benefit for many patients.^[4]^ Non-pharmacologic treatments included chiropractic therapy, physical therapy, massage therapy, and other interventions including ablative techniques, botulism toxin injections, electrical nerve stimulation, intrathecal drug therapies, epidural steroids, and minimally invasive spinal procedures.

### Outcome

Our primary outcome was a new OUD or opioid overdose diagnosis, defined as the presence of an ICD-10 diagnosis code for OUD,^[15,16,18]^ an ICD-10 diagnosis code for nonfatal, unintentional opioid overdose, or receipt of MOUD (methadone, buprenorphine, or extended-release injectable naltrexone).^[19]^ Outcomes were assessed during months 2-7 (i.e., after day 30 through month 7) and months 2-13 of follow-up for the primary analysis, and during months 4-9 and 4-15 for the sensitivity analyses.

### Statistical analysis

We estimated the extent to which *having versus not having each treatment modality* was associated with risk of a new-onset OUD or opioid overdose diagnosis, leaving other treatments in the set of 14 treatments as observed, and adjusting for measured covariates and censoring. For the opioid categories, comparisons were made by setting alternative opioid categories to not received while leaving nonopioid treatments as observed. To address potential violations of positivity, we also estimated effects comparing receipt of each treatment with the observed treatment pattern. In Section S2 of the Supplement, we show formal mathematical notation for these estimands and explain the assumptions necessary for these quantities to correspond to causal effects. Relative risks were estimated using a doubly robust, non-parametric, targeted minimum loss-based estimator with 2-fold cross-fitting.^[20]^ Exposure, outcome, and censoring models were estimated using an ensemble^[21]^ of machine learning algorithms, comprising an intercept-only model, a main-terms generalized linear model, multivariate adaptive regression splines^[22]^, gradient boosted machines,^[23]^ and least absolute shrinkage and selection operator (LASSO).^[24]^

Analyses were conducted using the *lmtp* package in R.^[25]^ All code to replicate analyses is available `blindedforreview`. This study was reported in accordance with the STROBE guidelines for observational research.^[26]^ This study was approved by the Columbia University Institutional Review Board.

## Results

The cohort included *N* = 525, 002 Medicaid patients who initiated treatment for newly diagnosed episodes of aLBP (Table 1). Most patients were women (61%), and 50% were non-Hispanic White patients. During the 6-month washout period, inpatient hospitalizations were rare (2.0%), while outpatient visits were very common (98.7% had ≥ 1 visit).

**Table 1:** Descriptive table for primary analysis.

| Characteristic | N=525,002 |
| --- | --- |
| **Baseline covariates (months -6 to 0)** |  |
| Age | 39 (29, 50) |
| Sex |  |
| Male | 203989 (38.9%) |
| Female | 321013 (61.1%) |
| Race/ethnicity: |  |
| White, non-Hispanic | 217871 (49.8%) |
| AIAN, non-Hispanic | 8129 (1.9%) |
| Asian, non-Hispanic | 34926 (8%) |
| Black, non-Hispanic | 78668 (18%) |
| Hawaiian/Pacific Islander | 3356 (0.8%) |
| Hispanic, all races | 94202 (21.5%) |
| Multiracial, non-Hispanic | 280 (0.1%) |
| Unknown | 87570 (16.7%) |
| Primary language English | 390326 (86.2%) |
| Unknown | 72373 (13.8%) |
| Married/partnered | 37495 (20.5%) |
| Unknown | 342121 (65.2%) |
| Household size: |  |
| 1 | 92806 (62.6%) |
| 2 | 19515 (13.2%) |
| ≥ 3 | 36030 (24.3%) |
| Unknown | 376651 (71.7%) |
| Veteran | 668 (0.6%) |
| Unknown | 411808 (78.4%) |
| High income (> 138FPL) | 13710 (2.6%) |
| TANF Benefits | 51445 (12.6%) |
| Unknown | 116694 (22.2%) |
| SSI benefits: |  |
| Mandatory or optional | 8142 (3.8%) |
| Not applicable | 207842 (96.2%) |
| Unknown | 309018 (58.9%) |
| Psychiatric conditions & counseling: |  |
| ADHD | 6974 (1.3%) |
| Anxiety | 65772 (12.5%) |
| Bipolar | 15423 (2.9%) |
| Depression | 49802 (9.5%) |
| Other mental illness | 24600 (4.7%) |
| Mental health counseling | 35201 (6.7%) |
| Alcohol use disorder | 12447 (2.4%) |
| Other drug use disorder (non-opioid) | 38214 (7.3%) |
| **Healthcare Utilization** |  |
| Emergency department |  |
| 0 | 389469 (74.2%) |
| 1-2 | 120201 (22.9%) |
| 3+ | 15332 (2.9%) |
| Inpatient hospitalizations |  |
| 0 | 514243 (98%) |
| 1+ | 10759 (2%) |
| Outpatient visits |  |
| 0 | 6788 (1.3%) |
| 1-25 | 498346 (94.9%) |
| 26+ | 19868 (3.8%) |

Table 1: continued
| Characteristic | N=525,002 |
| --- | --- |
| **Treatments (month 1)** |  |
| Acetaminophen | 26485 (5%) |
| Anti-inflammatory | 306165 (58.3%) |
| Benzodiazepine | 41915 (8%) |
| Chiropractic | 52973 (10.1%) |
| Duloxetine | 7292 (1.4%) |
| Gabapentin | 47529 (9.1%) |
| Intervention | 8742 (1.7%) |
| Muscle relaxant | 229522 (43.7%) |
| Massage therapy | 45355 (8.6%) |
| Physical therapy | 82997 (15.8%) |
| Spinal cord stimulation | 52 (0.01%) |
| Steroid | 81279 (15.5%) |
| Opioid | 106200 (20.2%) |
| ≤ 7 days, ≤ 50 MME | 67648 (12.9%) |
| > 7 days, ≤ 50 MME | 25758 (4.9%) |
| > 50 MME | 12794 (2.4%) |
| **Outcomes (months 2-13)** |  |
| OUD or overdose diagnosis by 7 months | 6553 (1.5%) |
| OUD or overdose diagnosis by 13 months | 8327 (2.4%) |
| **Censoring** |  |
| Uncensored through 7 months | 433777 (82.6%) |
| Uncensored through 13 months | 343434 (65.4%) |
| Binary values formatted as N (%). Continuous values formatted as median (IQR). |  |

Anti-inflammatory drugs (58.3%) and muscle relaxants (43.7%) were the most com-monly prescribed medications during the initial treatment period. Physical therapy was the most common non-pharmacologic treatment (15.8%). Among patients who filled an opioid prescription (20.2% overall), most received prescriptions for ≤ 50 maximum daily MME and ≤ 7 days of supply (64% of opioid recipients), while 12% received > 50 MME. The least common treatment, spinal cord stimulation (0.01%) was excluded from statistical analyses because of sparse data. Even for riskier treatments, like opioid prescriptions, the vast majority of beneficiaries received these prescriptions within the first few days of treatment. For example, out of all patients who received an opioid prescription within the first 3 months of treatment initiation, 75.9% had their opioid prescription start within the first week of treatment (Fig S1).

During months 2-13 of follow-up (Figure 4), gabapentinoids were associated with the largest increase in risk of new-onset diagnosis of OUD or overdose (increasing risk by 130.1%, 95% CI: 117.8%, 142.3%), followed by opioid dose exceeding 50 MME (118.1%, 95% CI: 99.2%, 137.0%). By contrast, opioid prescriptions ≤ 50 MME and ≤ 7 days were associated with a much smaller increase in risk of diagnosis of OUD or overdose (20.9%, 95% CI: 13.8%, 27.9%). Conversely, all non-pharmacologic, non-interventional therapies were estimated to be associated with reduced risk of new-onset diagnosis of OUD or overdose. Physical therapy was associated with the largest relative risk reduction (34.0% decrease, 95% CI: −40.9%, −27.1%). For months 2-7 of follow-up, chiropractic work was associated with the largest relative reduction (34.5% decrease, 95% CI: −41.4%, −27.7%), otherwise inferences remain unchanged (Figure 3). Estimates for absolute risk increases are provided in Table S3.

**Figure 3:**
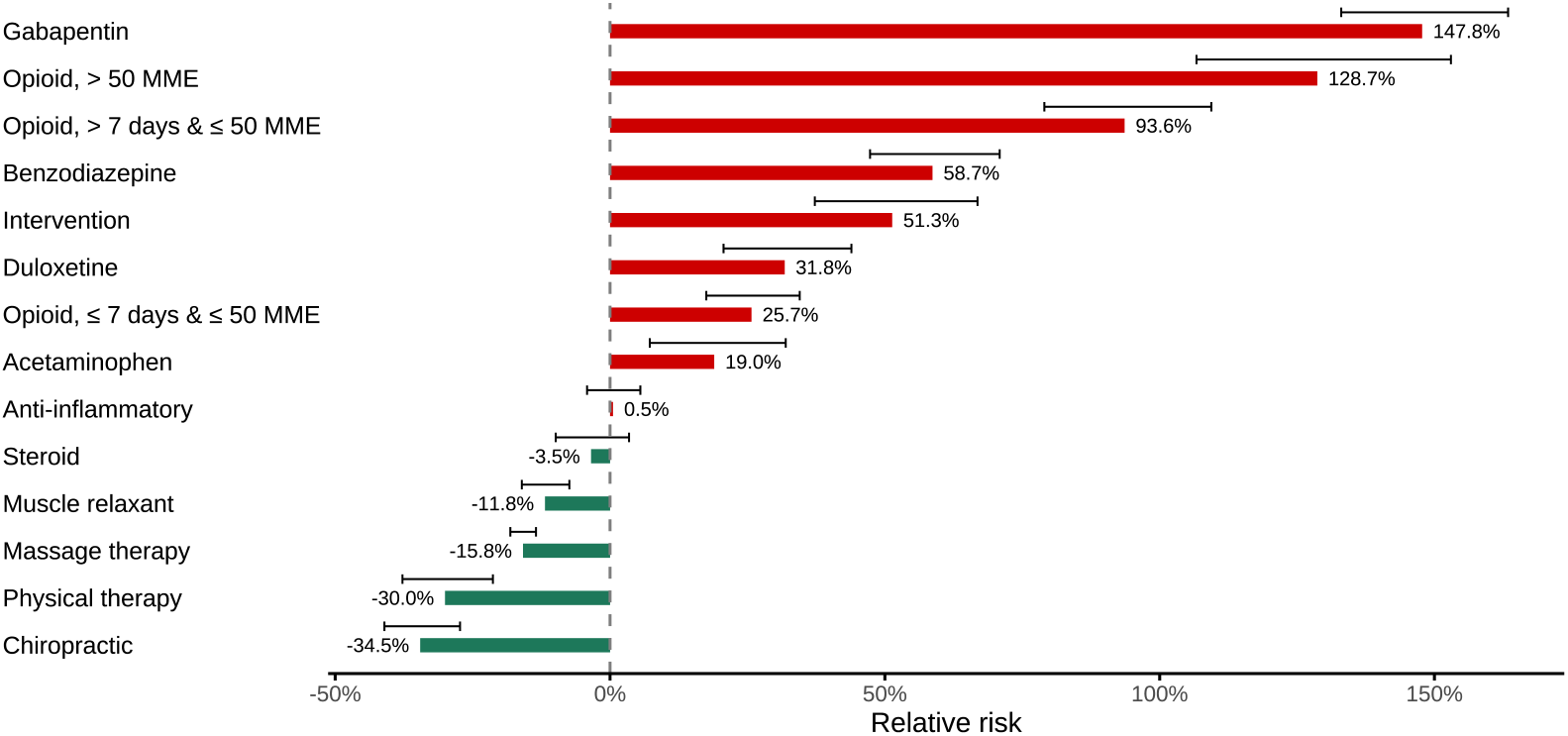
Low back pain patients with no previous OUD receiving a pain treatment within one month of diagnosis. Estimated relative risk of incident OUD over 6 months of follow-up hypothetically adding a treatment to initial treatment combination and no disenrollment, versus removing the treatment from initial treatment combination and no disenrollment. Error bars denote the 95% CI of the estimate.

**Figure 4:**
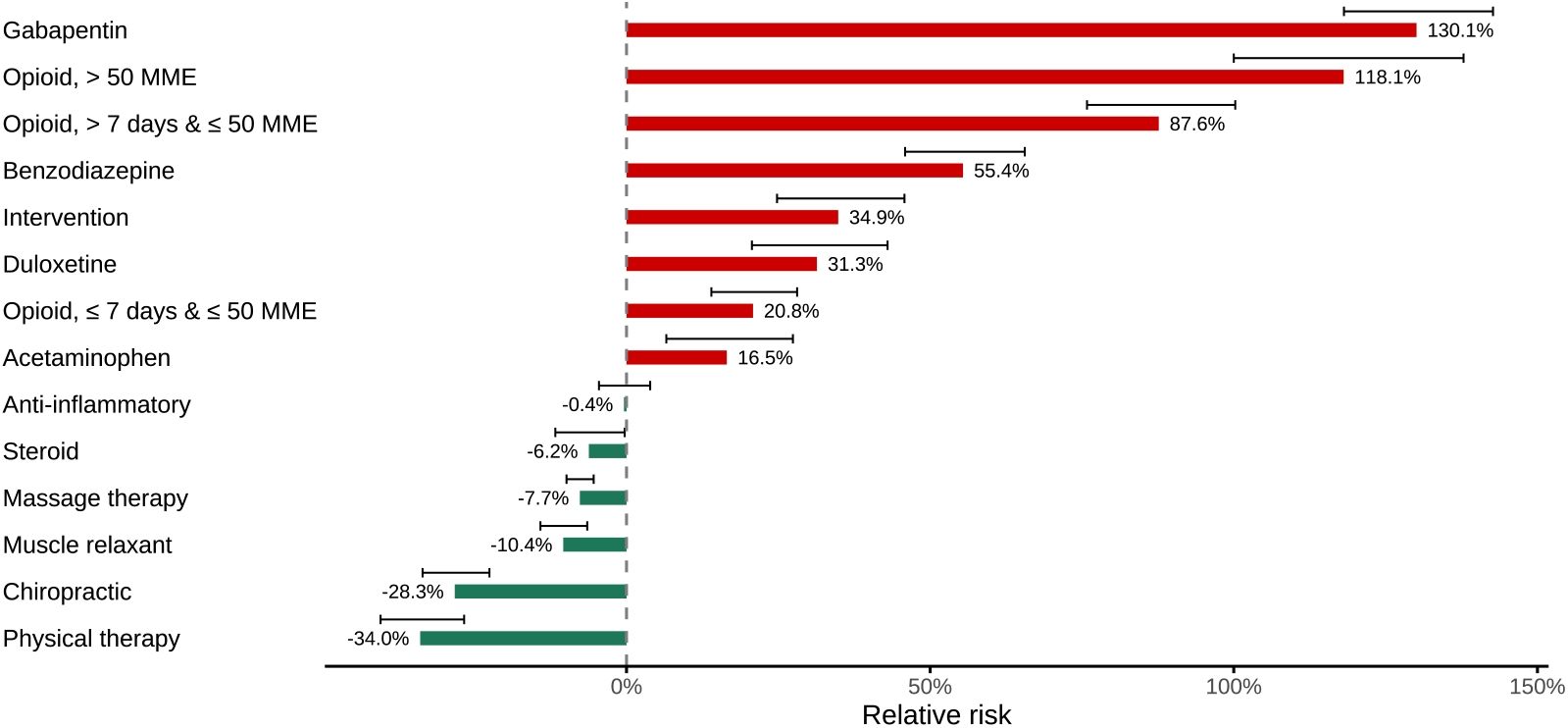
Low back pain patients with no previous OUD receiving a pain treatment within one month of diagnosis. Estimated relative risk of incident OUD over 12 months of follow-up hypothetically adding a treatment to initial treatment combination and no disenrollment, versus removing the treatment from initial treatment combination and no disenrollment. Error bars denote the 95% CI of the estimate.

As specified in the Methods section, we estimated associations for one treatment modality at a time, leaving the other treatments as observed. However, the frequency of co-prescribing may affect the average risks we estimate, with the co-prescription of gabapentinoids and opioids being particularly relevant. Table S2 shows the frequency their co-prescription, which was relatively rare in this cohort. Gabapentenoids were almost 4 times more likely to be prescribed *without* an overlapping opioid in the initial treatment period in this cohort, and opioids were more than 8 times more likely to be prescribed *without* an overlapping gabapentinoid.

Figures S2 and S3 show the estimated relative risks of new-onset diagnosis of OUD or overdose for each pain treatment versus its observed value. Relative to the with-versus-without-treatment comparison, the direction and ranking of relative risk remained consistent, although the effect sizes were attenuated, as expected. Most notably, lower dose, shorter duration opioid prescriptions resulted in minimal change in risk (9.1%, CI: 3.7%, 14.5%). Inferences were also robust in sensitivity analyses redefining the initial treatment episode as specified in the Methods section, using 30-day and 7-day allowable gaps between treatments for up to month 3 (Figures S4-S11 in the Supplement).

## Discussion

In this study of adult, opioid-naive Medicaid patients who initiated treatment for a new episode of aLBP, the cumulative incidence of new-onset diagnosis of OUD or opioid overdose was 1.5% and 2.4% at 7 and 13 months, respectively. Initial treatment patterns were predominantly pharmacological, most commonly anti-inflammatories, skeletal muscle relaxants, and opioids. Gabapentinoids and opioids were each associated with higher risk for a diagnosis of OUD or overdose. By contrast, non-pharmacologic, non-interventional treatment modalities were associated with risk reductions.

Consistent with prior studies, guideline-discordant care for aLBP was common. American College of Physicians guidelines for aLBP recommend non-pharmacologic therapies as first-line treatment;^[3]^ however, fewer than half of patients in this cohort received a nonpharmacologic therapy as part of initial care. Although anti-inflammatory medications and skeletal muscle relaxants, the recommended first-line pharmacologic options, were frequently prescribed, opioids were included in approximately 20% of initial treatment episodes and 17.3% of cohort patients received an opioid prescription within the first week of treatment.

The 2022 CDC Clinical Practice Guidelines for Prescribing Opioids for Pain state that opioid therapy for acute pain should only be considered when benefits are anticipated to outweigh risks.^[4]^ A recent randomized controlled trial cast doubt on the potential benefits of opioid therapy for acute low back or neck pain. This trial compared guideline-recommended care plus an opioid versus a placebo and found no difference in terms of pain severity at 6 weeks and a small but significant difference in pain severity at 1 year in favor of placebo. There were either no differences or small differences in favor of placebo for secondary outcomes of physical function, quality of life, and global perceived effect scale.^[27]^ These findings suggest that opioids should not be considered for aLBP. In our study, even low-dose, short-duration opioid prescriptions (≤ 50 MME and ≤ 7 days) were associated with a small increased risk of OUD or overdose diagnosis, with much higher risks associated with opioid prescriptions above 50 MME and/or duration ≥7 days. These findings are consistent with prior observational studies showing dose- and duration-dependent associations between opioid prescription and risk of OUD or overdose diagnoses.^[10,28,29]^

Gabapentinoids were associated with the highest risk of OUD or overdose diagnoses. Although sometimes perceived as safer alternatives to opioids, emerging evidence suggests important safety concerns. Among Medicare patients with aLBP, gabapentinoid prescribing has been associated with higher risk of drug-related overdose hospitalization within 90 days of aLBP diagnosis.^[9]^ Use of gabapentinoids in combination with opioids in other acute and chronic pain populations has been associated with respiratory depression and other opioid-related adverse events, including overdose, OUD, and death.^[10,30,31]^ Evidence supporting gabapentinoids for LBP is limited. A systematic review and meta-analysis of gabapentinoids for chronic LBP found that gabapentin did not provide significant pain improvement compared to placebo, pregabalin was less effective than other active comparators, and increased risk of adverse effects.^[32]^ Accordingly, guidelines for treatment of low back pain from the Department of Veterans Affairs and Department of Defense and guidelines from the National Institute for Health and Care Excellence do not recommend gabapentinoids for low back pain.^[33]^ In our cohort, gabapentinoids were included in approximately 10% of initial treatment episodes.

All non-pharmacologic, non-interventional therapies were associated with reductions in risk of overdose or OUD diagnosis, with physical therapy demonstrating the largest relative risk reduction. This is consistent with prior research showing associations between early physical therapy and reduced opioid use for treatment of aLBP and lower risk of long-term opioid use among patients with musculoskeletal pain.^[34,35]^ In a study of Medicaid patients with chronic pain, physical therapy was associated with the largest risk reduction in OUD out of a set of common pain treatments considered.^[10]^ In our cohort, physical therapy was included in only 16% of initial treatment episodes.

This study has several limitations. As an observational analysis of administrative claims data, results may be affected by residual confounding, misclassification, and unmeasured differences in pain severity and functional status. In terms of the outcome, although use of ICD-10 codes for opioid abuse or dependence in Medicaid claims have a sensitivity of nearly 90%, specificity is likely lower, and misclassification is a concern.^[36–38]^ For this reason, we refer to the outcome as OUD or overdose diagnosis. Over-the-counter medications and prescriptions covered by self-pay were not included (though self-pay is less common in the Medicaid population), and details on actual medication consumption including illicit use are unknown. The analysis did not account for all comorbidities or subtypes of aLBP that may influence treatment choices. Physician-directed exercise was also not captured. The trajectory of aLBP resolution or progression was unknown. It has been estimated that 2 to 48% of patients with aLBP transition to chronic low back pain, and some patients may have recurrent episodes of aLBP.^[39]^ Finally, findings may not generalize beyond the Medicaid population.

## Conclusions

For adults with newly diagnosed aLBP, initial treatment decisions may have meaningful downstream consequences. In this large Medicaid cohort, pharmacologic therapies, especially gabapentinoids and higher-dose opioid prescriptions, were associated with increased risk of subsequent diagnosis of OUD or overdose. In contrast, non-pharmacologic, non-interventional therapies, and physical therapy in particular, were associated with reductions in risk. These findings support prioritizing guideline-recommended non-pharmacologic, non-interventional therapies as first-line care for aLBP. Pharmacological therapies can be considered for carefully selected patients after weighing potential benefits against longer term risks. When opioids are prescribed, minimizing dose and duration remain prudent.^[4,5]^

## Data Availability

Data used in this study are proprietary and were obtained from the Centers for Medicare & Medicaid Services (CMS) Transformed Medicaid Statistical Information System (T-MSIS) analytic files. Data cannot be shared by the authors due to a Data Use Agreement.

