## supplement for "Associations between initial treatments for acute low back pain and opioid use disorder and overdose risk in Medicaid patients"

### S1 Additional detail on extracting TAF data

TAF data obtained for this study included the first 24 months of coverage for non-dual-eligible, non-pregnant, adult beneficiaries (ages 19-64), for whom Medicaid is the primary insurer, excluding eligibility group codes 05, 06, 07, 08, 23, 25, 29, 31, 33, 34, 44, 45, 53, 54, 61, 62, 64, 65, 66, and 68. Those with missing eligibility group codes were retained. Dual eligibility was defined as being non-dual-eligible for both Medicaid and Medicare for all enrolled months in the year. The years 2016-2019 and the following states were included: AZ, AR, CA, CO, CT, DE, HI, IA, IL, KY, MD, MA, MI, MN, NJ, NV, NH, NM, NY, ND, OH, OR, RI, VT, WA, WV. Additional subgroups were excluded, namely intellectual disability, blind/vision problem, deaf/hearing problem, and dementia/Alzheimer's. The 2016-2019 TAF Demographic and Eligibility files were searched for beneficiaries who meet all of the above criteria in the same year. The cohort was randomly reduced to 19,999,999 beneficiaries. For those beneficiaries, 2016-2019 TAF data were extracted in July 2022 for the Demographic and Eligibility, Other Services, Inpatient, and Pharmacy files.

### S2 Additional detail on statistical analysis

We assume observed data $\mathbf{O} = (\mathbf{W}, \mathbf{A}, C_1, C_1Y_1, C_2, C_2Y_2)$, where: $\mathbf{W}$ represents the covariates measured during the washout period; $\mathbf{A}$ represents initial pain management treatment variables; $C_t$ represents an indicator of remaining uncensored by follow-up time $t$, where $t = 1$ corresponds to month 9 (outcome measured during months 4-9) and $t = 2$ corresponds to month 15 (outcome measured during months 4-15); finally, $Y_t$ represents the outcome by time $t$, which is observed among those who remain uncensored.

We estimated two causal estimands. The first can be denoted

$$E(Y_t^{\mathrm{d1}_i, \bar{C}_t = \bar{1}} - Y_t^{\mathrm{d0}_i, \bar{C}_t = \bar{1}}), \tag{1}$$

where $\mathrm{d1}_i$ is defined as $\mathrm{d1}_i(\mathbf{A}) = (A_i = 1, \mathbf{A}\backslash A_i)$, $\mathrm{d0}_i$ is defined as $\mathrm{d0}_i(\mathbf{A}) = (A_i = 0, \mathbf{A}\backslash A_i)$, we use an overbar to denote variable history such that $\bar{C}_2 = \bar{1}$ denotes remaining uncensored at both follow-up timepoints, and we use the notation $Y^{\mathrm{d},\bar{1}}$ to indicate the counterfactual value of the outcome had the set of treatments been modified according to policy d and had censoring not occurred, possibly contrary to fact. The second causal estimand can be denoted

$$E(Y_t^{\mathrm{d1}_i,\bar{C}_t=\bar{1}} - Y_t^{\bar{C}_t=\bar{1}}), \tag{2}$$

where the second component of the contrast leaves the set of pain treatments as observed. $\mathrm{d1}_i$ and $\mathrm{d0}_i$ are defined as above for $i \in \{1, ..., 7, 12, 13, 14\}$. However, when hypothetically intervening on one of the three opioid variables, the other two variables must necessarily be set to 0. Consequently, for $i \in \{8, 9, 10\}, \mathrm{d1}_i(\mathbf{A}) = (A_i = 1, \mathbf{A} \in \{A_8, A_9, A_{10}\}\backslash A_i = (0, 0), \mathbf{A} \in \{A_1, ..., A_9, A_{10} \backslash A_i)$, $\mathrm{d0}_i$ is defined as $\mathrm{d0}_i(\mathbf{A}) = (A_i = 0, \mathbf{A} \in \{A_8, A_9, A_{10}\}\backslash A_i = (0, 0), \mathbf{A} \in \{A_1, ..., A_7, A_{11}, ..., A_{14}\}\backslash A_i)$.

Each of these causal estimands are equal to the following to statistical estimands, which are a function only of observed data and therefore estimable, under the identification assumptions given next:

$$E[E(Y_t \mid \bar{C}_t = \bar{1}, \mathbf{A} = \mathrm{d1}_i, \mathbf{W}) - E(Y_t \mid \bar{C}_t = \bar{1}, \mathbf{A} = \mathrm{d0}_i, \mathbf{W})], \tag{3}$$

$$E[E(Y_t \mid \bar{C}_t = \bar{1}, \mathbf{A} = \mathrm{d1}_i, \mathbf{W}) - E(Y_t \mid \bar{C}_t = \bar{1}, \mathbf{A}, \mathbf{W})]. \tag{4}$$

The identification assumptions under which the above statistical estimands equal the causal estimands are: 1) conditional exchangeability, meaning that there is no unobserved/unmeasured confounding of the relationship between the set of treatments and outcome conditional on covariates and that there is no unobserved/unmeasured confounding between censoring and the outcome conditional on the covariates and treatment; and 2) positivity, meaning that if it is possible to find a beneficiary with observed data $(\mathbf{a}, \bar{c}, \mathbf{w})$, then it is possible to find a beneficiary with data $(\mathrm{d1}_i(\mathbf{a}), \bar{1}, \mathbf{w})$ and $(\mathrm{d0}_i(\mathbf{a}), \bar{1}, \mathbf{w})$ for the first estimand pair and with data $(\mathrm{d1}_i(\mathbf{a}), \bar{1}, \mathbf{w})$ for the second estimand, for $i \in \{1:14\}$. In other words, this states that if it is possible to find a beneficiary with covariates $\mathbf{w}$, censoring $\bar{c}$, treatment set $\mathbf{a}$, it is possible to find a beneficiary with covariates $\mathbf{w}$, who are uncensored, and treatment set $\mathrm{d1}_i(\mathbf{a})$ and $\mathrm{d0}_i(\mathbf{a})$ for $i \in \{1:14\}$.

Table S1: International Classification of Diseases, Tenth Revision (ICD-10) codes used for assembling a cohort of individuals diagnosed with low back pain

| Pain category | ICD-10 code |
|---|---|
| Arthritis/Joint pain | M4005, M4015, M40205, M40295, M4035, M4036, M4037, M4045, M4046, M4047, M4055, M4056, M4057, M4105, M4106, M4107, M41115, M41116, M41117, M41125, M41126, M41127, M4125, M4126, M4127, M4135, M4145, M4146, M4147, M4155, M4156, M4157, M4205, M4206, M4207, M4215, M4216, M4217, M4305, M4306, M4307, M4315, M4316, M4317, M435X5, M435X6, M435X7, M438X5, M438X6, M438X7, M4625, M4626, M4627, M4635, M4636, M4637, M4845XA, M4846XA, M4847XA, M532X5, M532X6, M9903, M9983, S32000K, S32001K, S32002K, S32008K, S32009K, S32010K, S32011K, S32012K, S32018K, S32019K, S32020K, S32021K, S32022K, S32028K, S32029K, S32030K, S32031K, S32032K, S32038K, S32039K, S32040K, S32041K, S32042K, S32048K, S32049K, S32050K, S32051K, S32052K, S32058K, S32059K, S329XXK |
| Back Pain | M4325, M4326, M4327, M455, M456, M457, M4605, M4606, M4607, M4645, M4646, M4647, M4655, M4656, M4657, M4685, M4686, M4687, M4695, M4696, M4697, M4715, M4716, M4725, M4726, M4727, M47815, M47816, M47817, M47895, M47896, M47897, M4805, M48061, M4807, M4815, M4816, M4817, M4825, M4826, M4827, M4835, M4836, M4837, M488X5, M488X6, M488X7, M4985, M4986, M4987, M5105, M5106, M5115, M5116, M5117, M5125, M5126, M5127, M5135, M5136, M5137, M5145, M5146, M5147, M5185, M5186, M5187, M519, M532X7, M5385, M5386, M5387, M5405, M5406, M5407, M5415, M5416, M5417, M5430, M5431, M5432, M5440, M5441, M5442, M545, M9923, M9933, M9943, M9953, M9963, M9973 |
| Neurologic Pain | M47015, M47016, G541, G544 |

Figure S1: Days until opioid start among those with an opioid treatment (n=119,998). 91,087 individuals received opioids in first 7 days.

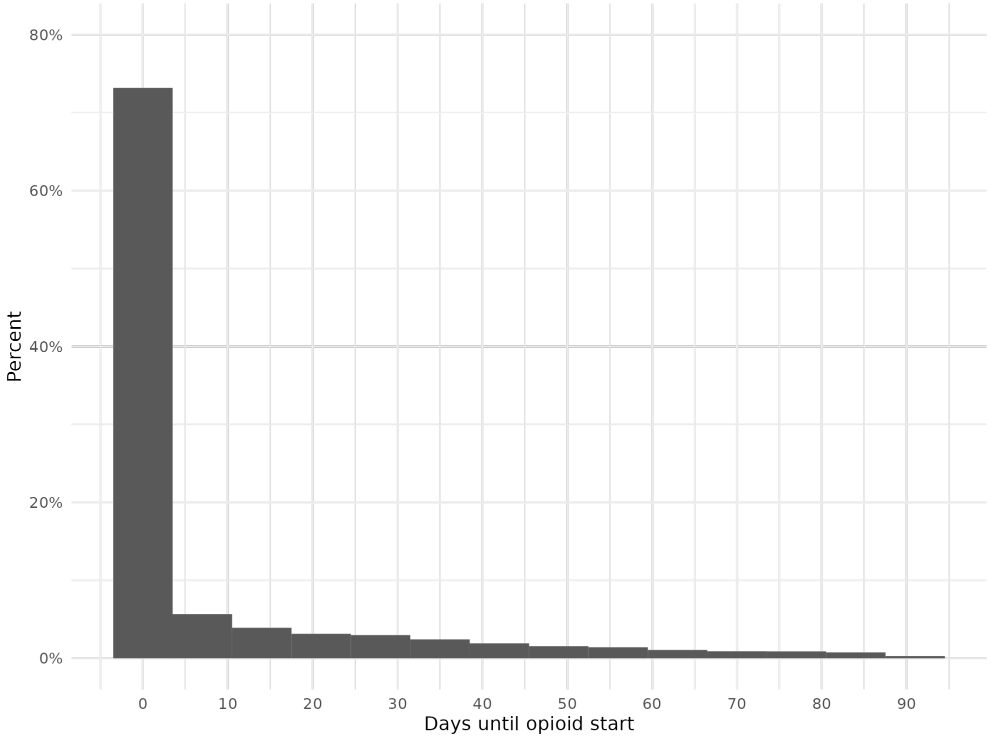


Table S2: Frequency of gabapentinoid and opioid exposure categories defined by overlap duration and opioid dose (N = 525,002).

| Co-prescription category | Number (%) |
|---|---|
| gabapentinoid *with* overlapping opioid $\leq$ 7 days, $\leq$ 50 MME | 3584 (0.7%) |
| gabapentinoid *with* overlapping opioid $>$ 50 MME | 2876 (0.5%) |
| gabapentinoid *with* overlapping opioid $>$ 7 days, $\leq$ 50 MME | 5849 (1.1%) |
| gabapentinoid *without* overlapping opioid | 42761 (8.1%) |
| opioid $\leq$ 7 days, $\leq$ 50 MME *without* overlapping gabapentinoid | 64097 (12.2%) |
| opioid $>$ 50 MME *without* overlapping gabapentinoid | 13405 (2.6%) |
| opioid $>$ 7 days, $\leq$ 50 MME *without* overlapping gabapentinoid | 25387 (4.8%) |

Binary values formatted as N (%).

Table S3: Low back pain patients with no previous OUD receiving a pain treatment within one month of diagnosis. Estimated risk differences of incident OUD hypothetically adding a treatment to initial treatment combination and no disenrollment versus removing the treatment from initial treatment combination and no disenrollment.

| Treatment | Risk difference at 6 months | Risk difference at 12 months |
|---|---|---|
| Gabapentin | 0.0192 | 0.0269 |
| Opioid, $>$ 50 MME | 0.0171 | 0.0249 |
| Opioid, $>$ 7 days & $\leq$ 50 MME | 0.0125 | 0.0185 |
| Benzodiazepine | 0.0082 | 0.0123 |
| Intervention | 0.0076 | 0.0081 |
| Duloxetine | 0.0047 | 0.0073 |
| Opioid, $\leq$ 7 days & $\leq$ 50 MME | 0.0034 | 0.0044 |
| Acetaminophen | 0.0028 | 0.0038 |
| Anti-inflammatory | 0.00008 | -0.0001 |
| Steroid | -0.0005 | -0.0015 |
| Massage therapy | -0.0024 | -0.0018 |
| Muscle relaxant | -0.0019 | -0.0026 |
| Chiropractic | -0.0054 | -0.0069 |
| Physical therapy | -0.0046 | -0.0083 |

Figure S2: Low back pain patients with no previous OUD receiving a pain treatment within one month of diagnosis. Estimated relative risk of incident OUD over 6 months of follow-up hypothetically adding a treatment to initial treatment combination and no disenrollment, versus holding treatments as observed and no disenrollment. Error bars denote the 95% CI of the estimate.

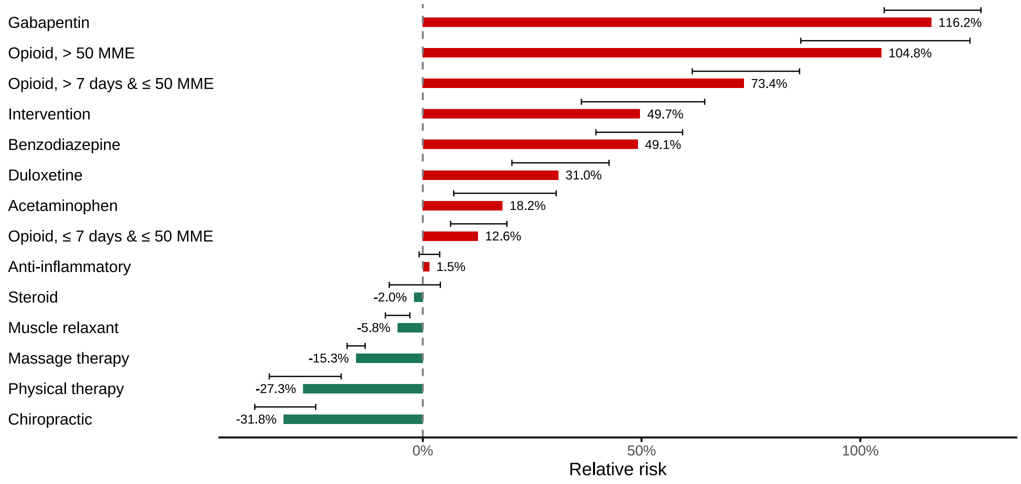

Figure S3: Low back pain patients with no previous OUD receiving a pain treatment within one month of diagnosis. Estimated relative risk of incident OUD over 12 months of follow-up hypothetically adding a treatment to initial treatment combination and no disenrollment, versus holding treatments as observed and no disenrollment. Error bars denote the 95% CI of the estimate.

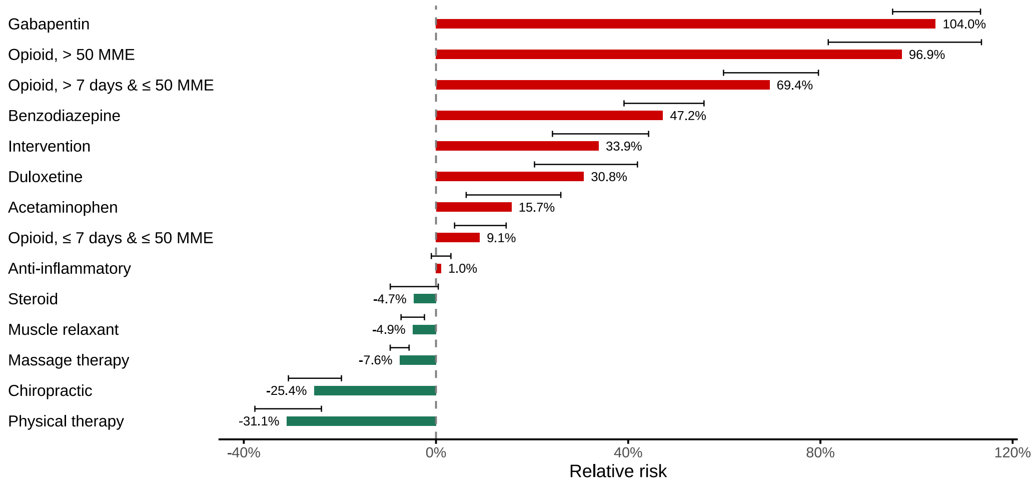

Table S4: Descriptive table for sensitivity analysis. Initial treatment period as ending at the first 30-day gap in pain treatment, or at the end of 3 months

| Characteristic | N=525,002 |
|---|---|
| **Treatments (months 1-3)** | |
| Acetaminophen | 30890 (5.9%) |
| Anti-inflammatory | 319849 (60.9%) |
| Benzodiazepine | 49226 (9.4%) |
| Chiropractic | 54471 (10.4%) |
| Duloxetine | 8920 (1.7%) |
| Gabapentin | 55070 (10.5%) |
| Intervention | 13426 (2.6%) |
| Muscle relaxant | 238559 (45.4%) |
| Massage therapy | 55078 (10.5%) |
| Physical therapy | 97712 (18.6%) |
| Spinal cord stimulation | 89 (0.02%) |
| Steroid | 93604 (17.8%) |
| Opioid | 119998 (22.9%) |
| $\leq$ 7 days, $\leq$ 50 MME | 70872 (13.5%) |
| $>$ 7 days, $\leq$ 50 MME | 32203 (6.1%) |
| $>$ 50 MME | 16923 (3.2%) |
| **Outcomes (months 4-15)** | |
| OUD or overdose by 9 months | 7402 (1.8%) |
| OUD or overdose by 15 months | 8631 (2.7%) |
| **Censoring** | |
| Uncensored through 9 months | 404253 (77.3%) |
| Uncensored through 15 months | 317394 (60.7%) |

Binary values formatted as N (%).

Table S5: Descriptive table for sensitivity analysis. Initial treatment period as ending at the first 7-day gap in pain treatment, or at the end of 3 months

| Characteristic | N=525,002 |
|---|---|
| **Treatments (months 1-3)** | |
| Acetaminophen | 26572 (5.1%) |
| Anti-inflammatory | 303111 (57.7%) |
| Benzodiazepine | 40647 (7.7%) |
| Chiropractic | 53033 (10.1%) |
| Duloxetine | 7245 (1.4%) |
| Gabapentin | 47047 (9%) |
| Intervention | 10630 (2%) |
| Muscle relaxant | 228797 (43.6%) |
| Massage therapy | 45816 (8.7%) |
| Physical therapy | 85663 (16.3%) |
| Spinal cord stimulation | 73 (0.01%) |
| Steroid | 82697 (15.8%) |
| Opioid | 107923 (20.6%) |
| $\leq$ 7 days, $\leq$ 50 MME | 66923 (12.7%) |
| $>$ 7 days, $\leq$ 50 MME | 26970 (5.1%) |
| $>$ 50 MME | 14030 (2.7%) |
| **Outcomes (months 4-15)** | |
| OUD or overdose by 9 months | 7402 (1.8%) |
| OUD or overdose by 15 months | 8631 (2.7%) |
| **Censoring** | |
| Uncensored through 9 months | 404253 (77.3%) |
| Uncensored through 15 months | 317394 (60.7%) |

Binary values formatted as N (%).

Figure S4: Sensitivity analysis redefining the initial treatment period as ending at the first 30-day gap in pain treatment, or at the end of 3 months. Estimated relative risk of incident OUD over 6 months of follow-up under hypothetically adding a treatment to initial treatment combination and no disenrollment, versus removing the treatment from initial treatment combination and no disenrollment. Error bars denote the 95% CI of the estimate.

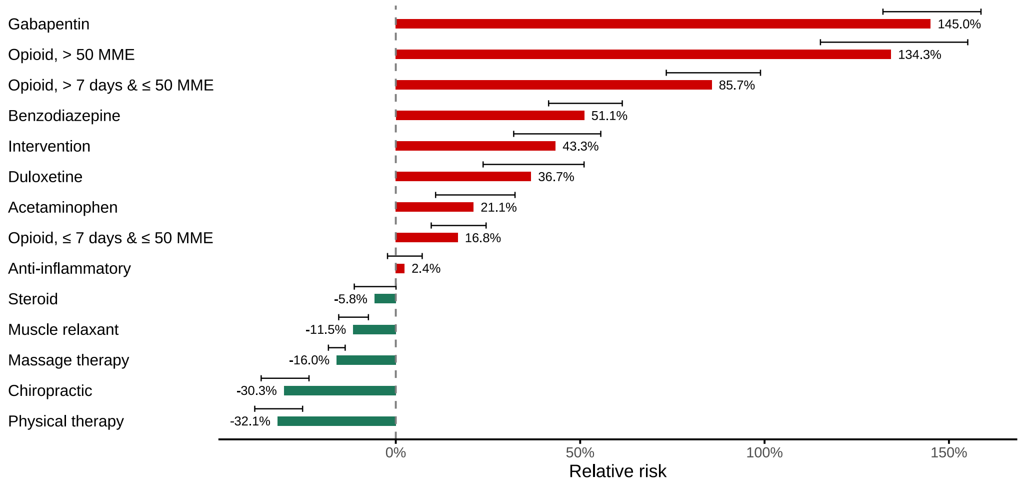


Figure S5: Sensitivity analysis redefining the initial treatment period as ending at the first 30-day gap in pain treatment, or at the end of 3 months. Estimated relative risk of incident OUD over 12 months of follow-up under hypothetically adding a treatment to initial treatment combination and no disenrollment, versus removing the treatment from initial treatment combination and no disenrollment. Error bars denote the 95% CI of the estimate.

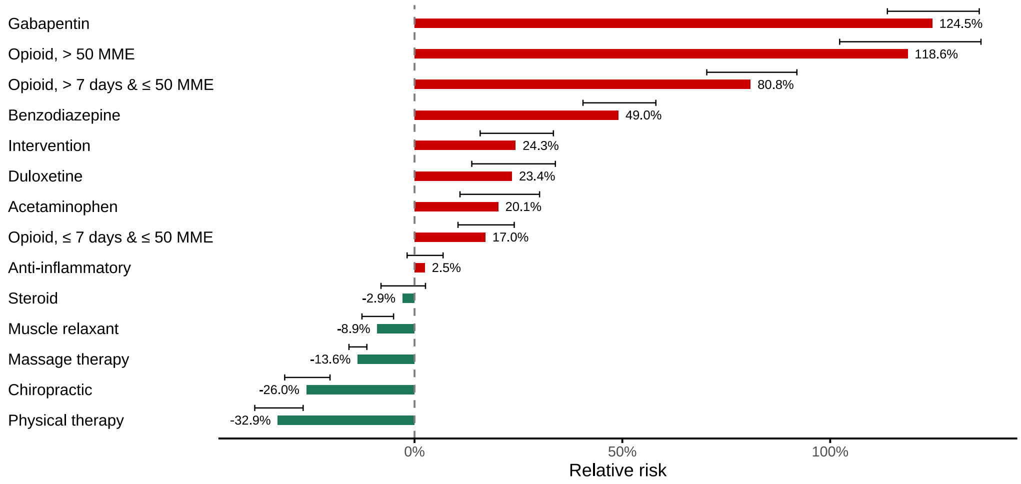

Figure S6: Sensitivity analysis redefining the initial treatment period as ending at the first 30-day gap in pain treatment, or at the end of 3 months. Estimated relative risk of incident OUD over 6 months of follow-up under hypothetically adding a treatment to initial treatment combination and no disenrollment, versus holding treatments as observed and no disenrollment. Error bars denote the 95% CI of the estimate.

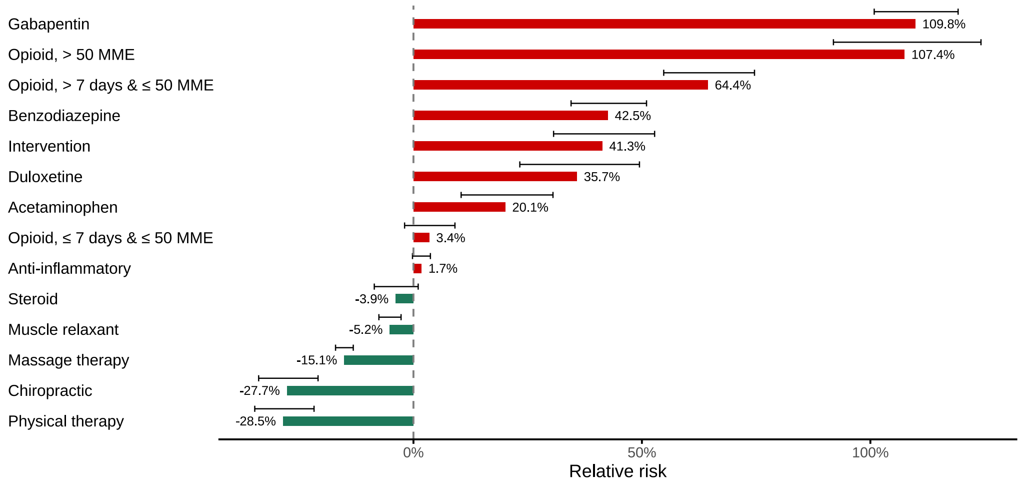


Figure S7: Sensitivity analysis redefining the initial treatment period as ending at the first 30-day gap in pain treatment, or at the end of 3 months. Estimated relative risk of incident OUD over 12 months of follow-up under hypothetically adding a treatment to initial treatment combination and no disenrollment, versus holding treatments as observed and no disenrollment. Error bars denote the 95% CI of the estimate.

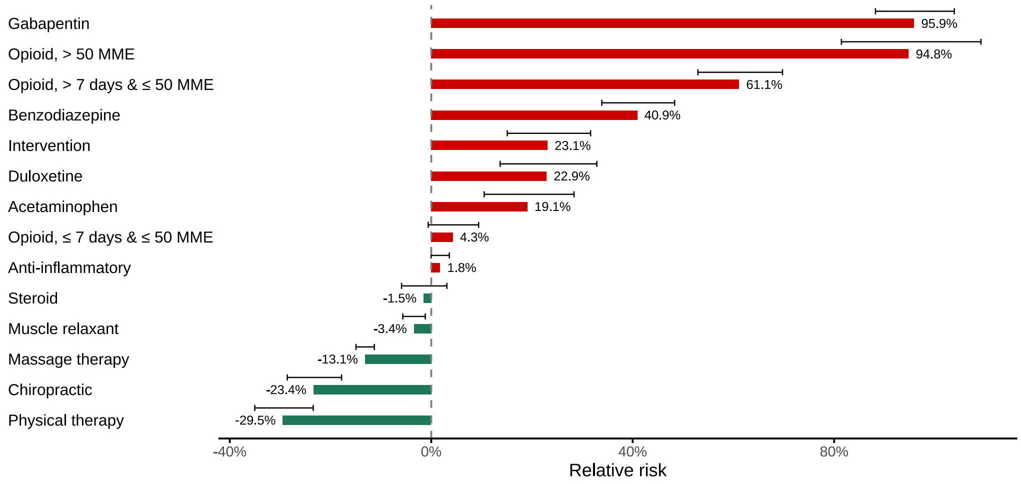

Figure S8: Sensitivity analysis redefining the initial treatment period as ending at the first 7-day gap in pain treatment, or at the end of 3 months. Estimated relative risk of incident OUD over 6 months of follow-up under hypothetically adding a treatment to initial treatment combination and no disenrollment, versus removing the treatment from initial treatment combination and no disenrollment. Error bars denote the 95% CI of the estimate.

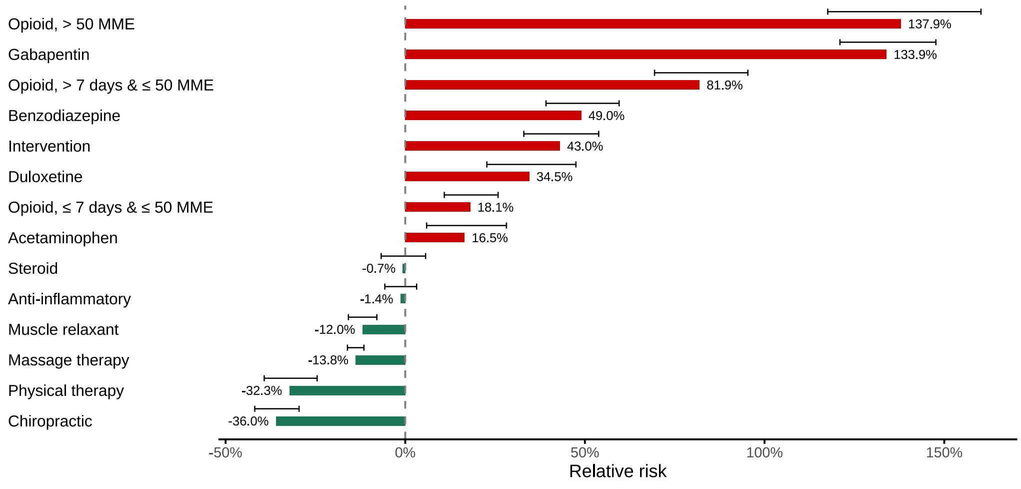


Figure S9: Sensitivity analysis redefining the initial treatment period as ending at the first 7-day gap in pain treatment, or at the end of 3 months. Estimated relative risk of incident OUD over 12 months of follow-up under hypothetically adding a treatment to initial treatment combination and no disenrollment, versus removing the treatment from initial treatment combination and no disenrollment. Error bars denote the 95% CI of the estimate.

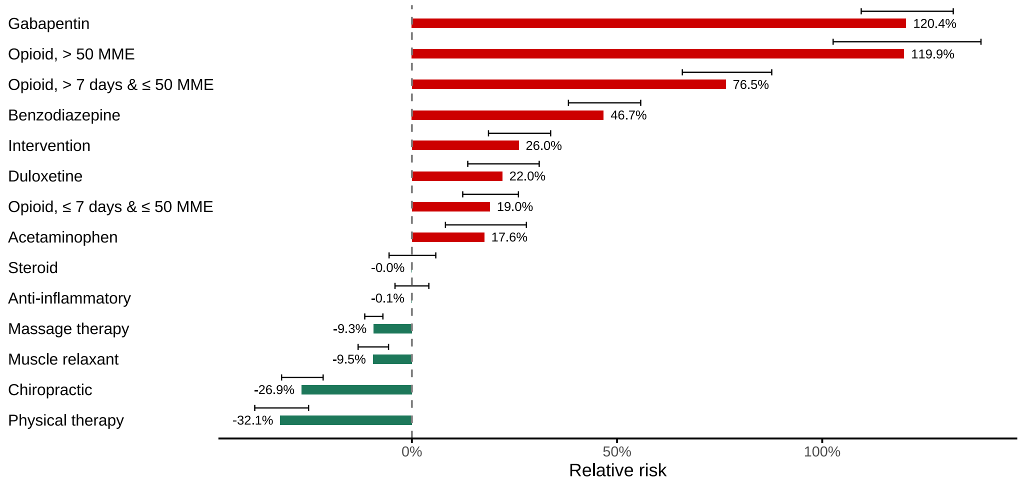

Figure S10: Sensitivity analysis redefining the initial treatment period as ending at the first 7-day gap in pain treatment, or at the end of 3 months. Estimated relative risk of incident OUD over 6 months of follow-up under hypothetically adding a treatment to initial treatment combination and no disenrollment, versus holding treatments as observed and no disenrollment. Error bars denote the 95% CI of the estimate.

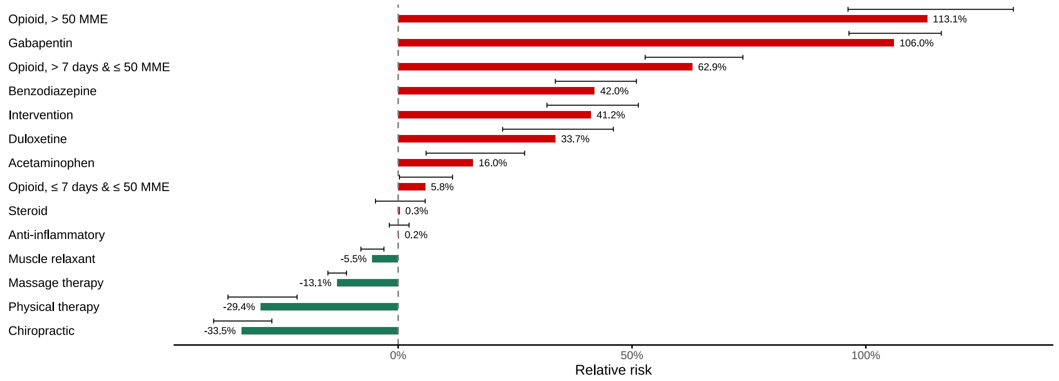


Figure S11: Sensitivity analysis redefining the initial treatment period as ending at the first 7-day gap in pain treatment, or at the end of 3 months. Estimated relative risk of incident OUD over 12 months of follow-up under hypothetically adding a treatment to initial treatment combination and no disenrollment, versus holding treatments as observed and no disenrollment. Error bars denote the 95% CI of the estimate.

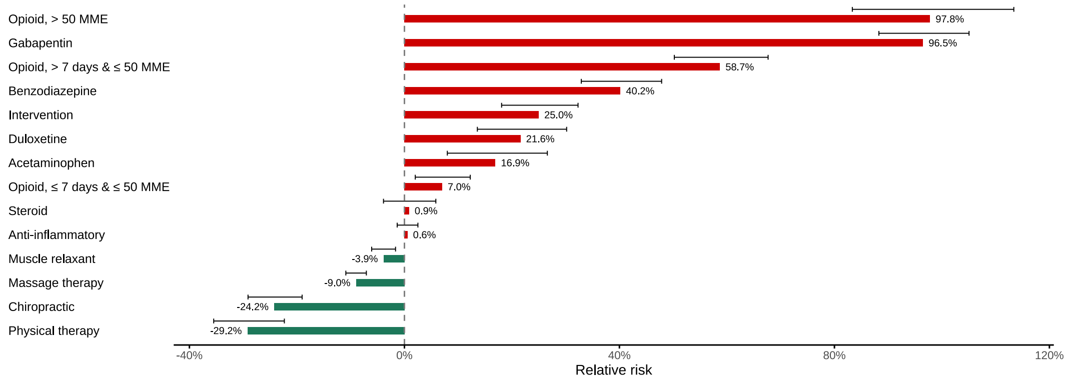
